# dqrep: A Stata package for automated data quality assessments and data monitoring

**DOI:** 10.1101/2025.07.06.25325294

**Authors:** Carsten Oliver Schmidt

**Author notes:** Corresponding author: Carsten Oliver Schmidt, Study of Health in Pomerania/KEF, University Medicine of Greifswald, Institute for Community Medicine, Walther Rathenau Str. 48, D-17475 Greifswald phone: +49[0]3834/86-7713 fax +49[0]3834/86-6684).

## Abstract

This article introduces `dqrep`, a Stata package designed for conducting comprehensive data quality assessments. A single command call flexibly scales from small “on-the-fly” assessments involving only a few variables to extensive tasks, such as generating and comparing quality reports for thousands of variables across multiple examinations within or across studies. To do so, `dqrep` activates an analytical pipeline that evaluates the requested data quality aspects, such as data integrity, missingness, range violations, outliers, temporal trends, observer or device effects. Detailed information and expectations about the data can be provided via the numerous `dqrep` options or MS Excel sheets. The package generates single or series of reports in PDF and DOCX formats, detailing data properties as well as the type, number, and severity of data quality issues. In addition, HTML dashboards may be requested to browse images. `dqrep` offers standardized machine-readable result summaries, facilitating downstream tasks such as benchmarking data quality across studies and examinations. The package is online available from https://dataquality.qihs.uni-greifswald.de/vignettes.html#STATA

## 1 Introduction

Data quality is commonly understood as data being “fit for purpose” (EMA Data Analytics and Methods Task Force, 2023). As such, it is a precondition for trustworthy research and requires thorough assessment. However, data quality reporting and initial data analysis (Huebner, le Cessie, Schmidt, & Vach, 2018) can be laborious and time-consuming despite several available software tools and statistics packages (Ehrlinger & Woss, 2022; Mariño, Kasbohm, Struckmann, Kapsner, & Schmidt, 2022). In addition, depending on the chosen approach, results may be difficult to integrate and compare across different application. Therefore, analysts would benefit from accessible analyses pipelines that enable a highly structured comprehensive, and standardized data assessment workflow. This would be particularly beneficial if the pipeline were able to trigger entire series of reports on multiple data sets, with subsequent benchmarking options to grade and compare data quality issues according to common standards.

With this in mind, `dqrep` was designed to facilitate complex data quality assessment tasks in observational health research. Examples are the population-based Study of Health in Pomerania (SHIP) (Volzke et al., 2022), the German National Cohort (GNC) (German National Cohort, 2014), euCanSHare (Devriendt et al., 2021), a joint EU-Canada project to facilitate cross-border data sharing based on a multi-cohort cardiovascular research platform, and the NFDI4Health (Pigeot et al., 2024), which focuses on epidemiological, public health and clinical research. However, the package is also of relevance to assess data in other fields of science.

The package takes into account the FAIR (Findable, Accessible, Interoperable, and Reusable) principles (Wilkinson et al., 2016). FAIRness is promoted on the input side through the use of machine-readable metadata tables, which specify reusable preconditions for data quality assessments. On the output side, in addition to advanced overviews of data properties and visualizations, standardized result files enable easy post-processing, such as benchmarking findings across examinations or studies.

## 2 The dqrep command

### 2.1 Syntax

`dqrep [varlist] [if] [in], [targetfiles(strings) sd(strings) lowercase(#) rd(strings) rdd(strings) gd(strings) ld(strings) replacereport(#) store(#) dashfile metadatafile(strings)`

sdmd(strings) hd(strings) not(varlist) gradingfile(strings) dataquieR keyvars(varlist) minorvars(varlist) processvars(varlist) controlvars(varlist) observervars(varlist) devicevars(varlist) centervars(varlist) timevars(varlist) case_order idvars(varlist) casemissvars(varlist) casemisstype(strings) casemisslogic(strings) varselect(varname) not(varlist) segmentname(varname) segmentselect(strings) segmentexclude(strings) benchmark(#) reportname(strings) reporttitle(strings) reportsubtitle(strings) authors(strings) reportformat(strings) pageformat(string) view_tp(#) view_integrity(#) view_dqi(#) view_changelog(#) dashboard(#) maxvarlabellength(#) varlinebreak sectionlinebreak linenumberpagebreak clustercolorpalettes(strings) decimals(#) heightadd(#) widthadd(#) language(strings) rename_ob(strings) rename_de(strings) rename_ce(strings) reporttemplate(strings) subgroup(strings) itemmisslist(numlist) itemjumplist(numlist) outcheck(#) outsens(#) outintegrate(#) binaryrecodelimit(#) metriclevels(#) minreportn(#) minvarnum(#) minclustersize_icc(#) minclustersize_lowess(#) minevent_count(#) minbwidth(#) histkat(#) problemvarreport(#) resultreport(#) nomod(#) forcecalc(#) breakreport(#)]

`dqrep` is designed to work with messy data and, therefore, does not assume any specific data properties a priori. This is necessary to avoid unwanted program terminations. Instead, it tries to recognize deficiencies and integrate them into a structured output that guides the users on further actions. `dqrep` operates in two distinct modes that mutually exclude each other:

1. First, `dqrep` can be used to assess variables in the active dataset by listing any variables of interest in `[varlist]`. This is useful for quick data inspections, although reports may also be complex when using the numerous `dqrep` options. In this mode, the working dataset is preserved, a copy is worked upon, modified data can be saved upon request and the original dataset is restored afterwards.
2. Second, `dqrep` can be used to perform checks of variables across an arbitrary number of datasets that still need to be loaded. These datasets can either be listed in the `targetfiles` option of the command or within a metadata Excel file (see section 4.3). This allows for a highly flexible application, integrating variables from different files. This enables full use of all `dqrep` functionalities. Specifying any variables in `[varlist]` before the comma will disable the second approach.

Currently, `dqrep` assumes that data from distinct files have at least partially overlapping common observational units.

To accommodate both approaches, the `dqrep` program call has no single mandatory component.

### 2.2 Options

`dqrep` offers a high degree of customizability through several groups of options:

#### 2.2.1 Options specific for the active dataset

Some options work with the active dataset only.

`ifin` Used to select cases of interest. For use with datasets that still need to be loaded, use the `subgroup` option instead.

`not`(varlist) Lists variables to be excluded from assessments.

`sort` If specified, the variables will be sorted alphabetically in the report.

#### 2.2.2 Target data files and folder options

These options concern the specification of data files and folders, as well as naming conventions.

`targetfiles`(strings) Contains the name of all data files to be analyzed. The “.dta” suffix can be omitted. The file names must not contain blanks. At least one file must be specified to run `dqrep` if an analysis is intended on variables other than those in the active dataset. If more than one name is specified, the files are merged 1:1 based on one or multiple specified key variables (option `idvars`). File names of interest can also be provided within the MS Excel metadata file. In this case, `targetfiles` should be left blank.

`sd`(strings) Source directory containing the Stata .dta files to be analyzed. More than one file may be used but all files must be stored in the same directory. If no directory is specified the current working directory is used.

`lowercase`(int) Whether or not to change variable names to lower letters (0=no, 1=yes). This option is useful to ensure an easier match between the names in the target data and metadata files. By default, lower letters are used if a metadata file is specified, otherwise no change is conducted. Check if this is appropriate.

#### 2.2.3 Result data files and folder options

Due to the potentially huge amount of output, there are multiple options to handle the storage of `dqrep` results. In case of complex reporting tasks, to improve oversight, it is recommended to store results in different directories. To function, `dqrep` requires rights to save files in the provided results directory or the active working directory.

`rd`(strings) Directory for any reports in PDF or DOCX format, as well as HTML gallery files.

`rdd`(strings) Result data directory to store tabular data quality result files and target data. This option is only of relevance if the store option has been specified. For more information on the result files check the store option. If nothing is specified, a subfolder “_dataresultfiles” is generated in the result directory “rd".

`gd`(strings) Result directory for graphical output. Graphical output is stored in an own folder because of the potentially large number of files. If nothing is specified, a subfolder “graph” is generated in “rd”.

`ld`(strings) Name of the directory for log files. If nothing is specified, log files are listed in “rd".

`replacereport`(`int 0`) Flag to replace an existing PDF or DOCX report: 0=no replacement, 1 always replace, 2 replace only PDF. If no replacement is chosen, the new report will be stored with ascending numeration, retaining the previous version.

`store(int 0)` Specifies whether auxiliary output should be saved in addition to a log file and report files. This comprises graphs, observational level target .dta files, and result files in .dta or XLSX format for postprocessing, as well as modified data in case of recoding due to range violations or outliers. The default 0, will save only the reports and log files, 1 will save all files except for XLSX files, and 2 all files including XLSX files.

`dashfile` Returns all results with a better readable variable naming to facilitate reuse outside the `dqrep` pipeline. In addition, the technical results file is also stored in a wide format.

#### 2.2.4 Metadata files and folder options

The `dqrep` package can handle information to control quality assessments by using Excel (.xlsx) files. This approach allows for a transparent set up and editing of the preconditions underlying quality reports. The permissible content of such files is described in section 4.3. The provision of a metadatafile is optional but strongly recommended to take full advantage of `dqrep`, please see examples.

`metadatafile`(strings) Contains the name of the metadata file that provides the information to control data quality analyses. The expected format is Excel (.xlsx).

`sdmd`(strings) Directory where the metadata file is stored. If nothing is specified, either the source directory folder `sd` is used or the current working directory.

`hd`(strings) Name of the directory containing help files to further control the report generation such as a grading rule files. If nothing is specified either `sdmd, sd` or the current working directory is used, depending on what is available.

`gradingfile`(strings) Specifies the name of an Excel xlsx file that contains the rules and output formats for data quality gradings. Any rules that have been applied to the data are listed in the output. The Excel grading template file example_grading.xlsx contains an explanation on how to set up grading rules.

`select_dqi`(strings) Specifies which indicators to apply when loading a grading file. This is useful to limit gradings to specific aspects of interest. If not provided, all rules will be applied. A list of of grading rule IDs (column ind_link in the grading file) must be specified, separated by a blank, e.g. `select_dqi(ID01 ID05 ID10)`. The meaning is as follows: I01 Maximum data quality problem; I02 Item missing in Percent (not taking into account user specified missing reason); I03 Item Response in Percent (taking into account user specified missing reason); I04 Proportion of values per variable violating inadmissibility limits (hard limits); I05 Proportion of values per variable violating uncertainty limits (soft limits); I06 Proportion of values identified as univariate outliers; I07 Variance proportion due to Level 2 cluster variable based on a fixed effects model; I08 Variance proportion due to Level 2 cluster variable based on a random effects model; I09 Variance proportion due to Level 3 cluster variable based on a random effects model; I10 Variance proportion due to Level 2 cluster variable based on a random effects model; I11 Variance proportion due to time trends.

`dataquieR` If dataquieR is specified, dqrep assumes that metadata is delivered in the format of the R dataquieR package (Struckmann, Marino, Kasbohm, Salogni, & Schmidt, 2024), which can be downloaded from CRAN. Note that only those dataquieR columns will be used that have a correspondence in dqrep metadata scheme as detailed below.

#### 2.2.5 Variable selection and variable role options

The following options provide a means to select variables into distinct roles and by doing so control their handling during assessments. The `reporttemplate` option makes use of these roles to assign distinct analysis options. The output in the result tables of the quality reports is grouped by variable role. Some roles directly influence assessments, such as specified cluster variables or time variables. Such roles take on a global meaning, for instance, the same cluster variable applies across all key variables. Variable roles can also be assigned through the metadata file, enabling a more specific assignment, e.g. cluster variables specific to each key variable. This considerable strengthens the adaptability of reports. A single variable can only be processed in one assigned role. If erroneously specified multiple times, the assumed most important role gets assigned.

`keyvars`(varlist) List of the high-priority variables for data quality assessments. For these, the most extensive computations take place. Commonly each key variable receives a dedicated output page with graphs. Not mentioning ‘keyvars’ leads to a use of all variables as key variables unless they have been assigned to another variable category.

`minorvars`(varlist) List of lower-priority variables, for which a more concise scope of data quality assessments is conducted. Typically, each variable in this category is provided with only a tabular overview.

`processvars`(varlist) These variables relate to process aspects of data, such as date-time stamps or ambient conditions. Typically, they play no role as outcome variables.

`controlvars`(varlist) A list of control variables to adjust effects in regression analyses, currently focusing on determining cluster effects.

`observervars`(varlist) A list of categorical variables intended to analyze observer effects. Any variable here is referred to as observer in the output. Because referring to observers may not be adequate in a range of applications, the output name may be altered with the `rename_ob` option.

`devicevars`(varlist) A list of categorical variables intended to analyze device effects. Any variable here is referred to as device in the output. Because referring to devices may not be adequate in a range of applications, the output name may be altered with the `rename_de` option.

`centervars`(varlist) A list of categorical variables intended to analyze observer effects. Any variable here is referred to as center in the output. `centervars` may be treated as a third level in a hierarchical multilevel analysis. Because referring to centers may not be adequate in a range of applications, the output name may be altered with the `rename_ce` option.

`timevars`(varname) May contain one date-time stamp to analyze time trends. `dqrep` tries to automatically recognize the format of date / time stamps, if not provided adequately.

`case_order` If no time variable is available, the order of cases in the file can sometimes be useful for assessing trends in the data. Specifying `case_order` enables this.

`idvars`(varlist) A list of variables used to create an ID variable that uniquely identifies each row in the dataset, to detect duplicates and to enable the merging of two or more datasets. Unintended non-uniqueness will result in merging failures. Users will be informed about this in the integrity output.

`casemissvars`(varname) Name of the variable used to define unit missingness, meaning observational units that did not enter the data collection.

`casemisstype`(strings) This option is used to describe the meaning of the variable that describes meaning of the case selection in the output based on casemissvars.

`casemisslogics` (strings) Specifies the logic to identify available observations (e.g. <50) using casemissvars.

`varselect`(varname) This option is relevant only when using a metadata (.xlsx) file. It specifies an identifier variable in a metadata file that defines which variables are to be included in a report. The specified variable in the metadata file must marked as ’1’ for it to be included in the report. Any other value is ignored. This option is particularly useful when targeting only a subset of the variables in a metadata file for inclusion in a report. Another option for groups of variables is the `segmentselect` option in the following paragraph.

#### 2.2.6 Multi reporting and benchmarking options

Structuring reports in a reasonable manner—such as by examination, data collection method, or other criteria can be useful gain a better oversight in case of larger variable numbers. The options in this section enable the sub-setting of variables into different reports to enhance such oversight. All following options are applied only when using an .XLSX metadata file. Additionally, it can be of interest to compare results across reports. This is controlled with the benchmarking option.

`segmentname`(varname) Specifies the name of the variable in the metadata file used to define segments. Ideally, this should be a string variable containing a readable name for each group of variables. All variables belonging to the same segment will be included in the same report. By default, each unique string within segmentname is treated as one segment. Segment names must not contain blanks.

`segmentselect`(strings) Sometimes, only a subset of segments from `segmentname` is relevant. A subset can be specified using this option, with each name separated by a space. `segmentselect` and `segmentexclude` can jointly be used.

`segmentexclude`(strings) If many segments are defined in the metadata file and only few are to be excluded, this option can be used. In this case all segments but the excluded one will be targeted with one report per segment. `segmentselect` and `segmentexclude` can jointly be used.

`benchmark`(int 0) When generating multiple reports based on `segmentname`, specifying the benchmark option creates an additional report to benchmark results across all computed reports, based on the applied data quality grading. The value specified in `benchmark` indicates the minimum problem level a variable must reach to be included in the data quality overview table, to enable a better focus on issues of interest. The default value ‘0’ suppresses benchmarking. Benchmarking output is also suppressed when disabling quality gradings.

#### 2.2.7 Report formatting options

There are numerous options to customize the formatting of data quality reports to satisfy users’ needs. These include specifications for report names, authors, and output formats, as well as more detailed adjustments such as managing page breaks or modifying graph sizes to improve layout.

`reportname`(strings) Defines the name used to store the data quality report. This name should be short and concise, without spaces. If spaces are present, they will be replaced by “_”.

`reporttitle`(strings) Defines the title of the report to be displayed as the header in output documents.

`reportsubtitle`(strings) Defines the subtitle of the report to be displayed as the subheader in output documents.

`authors`(strings) The authors of the report. These appear on the title page below the subheader.

`reportformat`(strings) Specifies the output format of the report, which can be either PDF (default) or DOCX. The DOCX format facilitates editing.

`pageformat`(strings) Specifies the page size for PDF or DOCX output. Permissible options are “letter” or “A4.” If no option is specified, the default is “letter” for English output and “A4” for German output.

`view_tp`(int) By default, no title page is displayed for descriptive overviews, while a title page is included for other report types. This option allows users to override the default setting, with ‘0’ suppressing the title page and ‘1’ requesting it.

`view_integrity`(int) By default, full data quality reports include an overview table addressing structural or technical issues, such as unexpected data types or non-unique IDs.

`view_integrity` allows users to override the default setting. (0=no integrity section, 1=display integrity section). For descriptive reports this output is suppressed by default.

`view_dqi`(int) By default, only the templates “standard, “extended, and their variations result in reports that include a grading of data quality issues. `view_dqi` allows users to override the default setting. (0=no data quality grading section, 1=display data quality grading)

`view_changelog`(int) By default, data quality reports include a change log with all conducted variable modifications. `view_changelog` allows users to override the default setting. (0=no change log, 1=display change log). For descriptive reports this output is suppressed by default.

`dashboard`(int 0) Generates an HTML gallery in the result folder to review all created images, when set to 1 or 2. This feature is particularly useful for datasets with a large number of variables, as it provides an accessible overview. Images can be resized and searched by the file name, which includes the variable name. The images in the dashboard contain links to itself to view and compare selections in a resizable pop-up window. Setting `dashboard` to “2” disables PDF or DOCX reporting entirely. Activating the dashboard automatically retains all stored result files. It is recommendable to define a dedicated result directory for this purpose.

`maxvarlabellength`(int 50) Variable labels may be too long for a readable report output. This option can therefore be used to limit the length of variable labels.

`varlinebreak`(int 1) Specifies whether a page break occurs after presenting results for each variable in the single-variable section of a report. Selecting “no” may be preferable for descriptive outputs. (0=no, 1=yes:default)

`sectionlinebreak`(int 1) Specifies whether or not a page break occurs after each summary table and report section. (0=no, 1=yes:default)

`linenumberpagebreak`(int 7) For few variables more than one table may be presented on a single page. This option specifies the number of rows required for a table to accept a page break afterwards and overrides the previous option. (default is n=7 rows in a table, before a page break occurs.)

`clustercolorpalettes`(strings) Specifies a list of color palettes to be assigned to graphical elements related to clusters (e.g., observers or devices). The first palette is assigned to the first cluster (e.g., examiners, devices), the second to the second cluster, and so on. If only one color is specified, its intensity is graded according to the number of clusters. If there are more clusters than palettes, the last listed palette is used for all remaining clusters. The current default palettes are “s1 economist s2 burd s1r s2 plottig".

`decimals`(#) Number of decimals to be displayed in output tables (default n=2).

`heightadd`(int 0) Adds a constant to adjust the height of graphs (default: *n* = 0). This can be important for enabling page breaks at desired positions in the single-variable output. Another use is creating larger graphs for use outside PDF or DOCX reports.

`widthadd`(int 0) Adds a constant to adjust the width of graphs (default: *n* = 0). This can be important for enabling page breaks at desired positions in the single-variable output. Another use is creating larger graphs for use outside PDF or DOCX reports.

`language`(strings) Specifies the output text for the PDF or DOCX reports (Default is e=English; d=German/deutsch). The log file language is English only. Other languages may easily be added upon request.

`rename_ob`(strings) Replaces the default output for `observervars` by a user defined string to improve report readability. Example: `observervars(batch) rename_ob(batchnumber)`will change the PDF or DOCX output to display “batchnumber” instead of “observer” in applicable tables and graphs.

`rename_de`(strings) Replaces the default output for `devicevars` by a user defined string to improve report readability.

`rename_ce` (strings) Replaces default output for `centervars` by a user defined string to improve report readability.

#### 2.2.8 Analysis options

Analysis options control various computational aspects of the report generation. The first aspect involves specifying the scope of the intended analyses, such as whether the report should focus on specific issues like missing data. The second aspect tailors the scope of analyses to different variable categories. The third aspect controls details of how analyses are conducted, including methods used to assess outliers or minimum case requirements to perform analyses.

`reporttemplate`(strings) This option is the primary control for defining the scope of data quality reports. If nothing is specified, one of two default settings is applied: for a dataset without a metadata file, dqrep assumes a simple descriptive overview is needed; with a metadata file, a “standard” data quality report is generated, including tables, graphs and a grading of data quality issues. If there is an explicit differentiation of key variables as well as process variables via command call options, a standard report will be generated without quality grading. Any default can be overridden by requesting one or more of the following templates: “D": Brief descriptive statistics with graph; “D2": Descriptive statistics without graph; “D3": Extended descriptive statistics with graph; “M": Missingness only; “RV": Range violations; “AC": Accuracy (outliers, cluster effects, time trends); “UO": Univariate outliers; “TREND": Assesses (time) trends using lowess regression; “VP2": Variance proportion in two level structure; “VP3": Variance proportion in three level structure; “tables": report with tables only; “standard": Normal report scope with detailed information on key variables and a table overview for other variable categories “extended": Standard report plus detailed coverage of minor and process variables. Both, “standard” and “extended” can also be entered as “standard3” and “extended3". This computes variance components in a hierarchical three level design situation, e.g. observers at level 2 nested in centers at level three. The third level cluster variable must be provided with the `centervars` option. Appending a “+” to the full report options (e.g., “standard+") will add graphs on absolute and relative frequencies for categorical outcomes and violin plots for continuous outcomes.

`subgroup`(strings) Defines the logic to make a subgroup selection. For example, if only persons younger than 30 years of age are to be selected the command call would be subgroup(age<30). `itemmisslist`(numlist) A list of numerical values to be treated as missing values, meaning expected values have not been encountered. The numerical lists may contain Stata missing codes (e.g. “.j .z”). A numerical list should contain at least one value. There are several options to simplify the input: 90/99 -> “91 92 93 94 95 96 97 98 99” / 900(10)990 -> “900 910 920 930 940 950 960 970 980” / 8(.5)10 90 91 96/99 -> “8 8.5 9 9.5 10 90 91 96 97 98 99”. Information from `itemmisslist` is ignored if respective metadata information from an XLSX sheet can be used. `itemjumplist`(numlist) A list of numerical values to be treated as permitted jumps (value not encountered and not expected, for example value not collected due to study design such as number of pregnancies in males). The same formatting rules apply as for `itemmisslist`.

Information from `itemjumplist` is ignored if respective metadata information can be used. `outcheck`(int 1) Specifies the type of outlier checks to be applied: ‘1’: Medcouple and Grubbs test (default, chosen to combine strengths for symmetric and asymmetric distributions.); ‘2’: All checks except Tukey; ‘3’: All checks including Tukey; ‘10’: only Medcouple (Rule according to G. Bray et al. (2005)); ‘11’: only Standard deviation based (default 3*SD); ‘12’: only Grubbs Test (default CI level 95); ‘13’: only Adjusted Tukey (default p10-2*(p25-p5) / p90+2*(p95-75)); ‘19’: only Tukey. How to integrate findings is specified using the `outintegrate` option.

`outsens`(real 1) Adjusts the sensitivity of outlier checks. The default value is 1, indicating no change. Increasing this value increases the threshold by the chosen factor for all applicable approaches (e.g., entering 1.5 changes the default for the Standard deviation based approach from 3 SD to 4.5 SD).

`outintegrate`(int 1) Determines how results from different outlier detection methods are integrated. The following options exist: ‘1‘: Adaptive approach with Medcouple and Grubbs (default). Both must indicate an outlier on the longer tail of the distribution, only Medcouple must indicate an outlier on the shorter tail; ‘2‘: All selected checks must indicate a value as an outlier; ‘10‘: only Adjusted boxplot with Medcouple; ‘11‘: only Standard deviation based approach; ‘12‘: only Grubbs Test; ‘13‘: only Adjusted Tukey; only ‘19‘ :Tukey. Rather strict default settings are used because the interest is foremost in error outliers not influential observations per se. The importance of these tests is considerably reduced if ranges for inadmissible or uncertain values are specified in the metadata file.

`binaryrecodelimit`(int 8) For nominal variables, assessing marginal effects or displaying trends across all levels of an outcome variable easily results in unreadable or excessive output. By default, such variables are recoded to binary to provide an initial insight into potential effects. This option controls the number of value levels up to which recoding to a binary variable should occur in the absence of a metadata file. If set to ‘0,’ binary recoding is suppressed entirely. The most frequent category is chosen as default. To avoid issues with nonsensical recodings, it is recommended to define such rules in the metadata XLSX file. If an assessment of clusters is desired for all levels of a categorical variable, it should be provided to `dqrep` as dummy-coded. No recoding (0) is the default for descriptive reports, 8 is the default for other report types.

`metriclevels`(int 25) Specifies the number of categories beyond which a variable is classified as being of interval or ratio scale type if no such information is available in the metadata file. This is a potentially error-prone heuristic, so full use of the XLSX metadata attribute `scalelevel` is strongly recommended.

`minreportn`(int 30) If the number of cases is too low, generating a data quality report may not be meaningful. This option specifies the minimum number of cases required to generate a report, with the default set to *N* = 30.

`minvarnum`(int 1) Minimum number of valid variables to generate a report, with the default set to N=1. A higher number may be specified to avoid nuisance reports.

`minclustersize_icc`(int 20) If the number of cases is too small, generating reliable cluster effects may not be feasible. This option specifies the minimum cluster size required to compute ICC values, with the default set to *n* = 10. Clusters with fewer cases are excluded from the computation.

`minclustersize_lowess`(n 40) If the number of cases is too small, assessing reliable trends may not be feasible. This option specifies the case number within clusters to compute time trends based using LOWESS regressions, with the default set to *n* = 40. Clusters with fewer cases are excluded from the computation.

`minevent_count`(int 5) Minimum number of events on the outcome variable to compute a logistics regression, with the default set to n=3.

`minbwidth`(real 0.2) To control the minimum permissible bandwidth in lowess analyses for trends.

`histkat`(int 15) Specifies the maximum number of value levels for which a bar chart is used when no additional information about the scale level of a variable is available. If the number of levels exceeds this limit, a histogram is used instead. (Default is n=15)

`problemvarreport`(#) Produces an additional report which contains an in depth analysis of all variables assigned to an issue category n=# or higher based on the data quality grading. This request makes sense in case of reports containing many variables to get a focused overview on variables with potential issues. The number is the minimum graded data quality category with the default options being set to 1 “OK”; 2 “uncertain”; 3 “Note”; 4 “low”; 5 “moderate”; 6 “important”; 7 “critical”. For an explanation of the levels check the chapter on quality grading.

`resultreport`(int 0) If a data quality report is to be generated from already existing `dqrep` result files, the option `resultreport` must be set to 1. If only a benchmark report is to be created based on a set of existing result files, the option `resultreport` must be set to 2. In the latter case, regular data quality reports will be not be generated, only the benchmark report is generated. This is important to save computational time. In both cases, all relevant dqrep report files must be provided with the option `targetfiles`.

`nomod`(int 0) This option controls `dqrep` data management procedures such as the deletion of detected inadmissible values, uncertain values, and outliers. When such values are detected, they are deleted to avoid counting the same data quality issue across different indicators. However, in specific circumstances this may not be desirable. Modifications can therefore be fine-tuned at the price of losing clarity of quality assessments. The options are: 0 ’All modifications permitted’ (default); 1 ’Modifications only permitted for inadmissible values and uncertain values’; 2 ’Modifications only permitted for inadmissible values’; 3 ’Modifications only permitted for outliers’. 4 ’No modifications permitted’; All conducted modifications are listed in the change log. No change (4) is the default for descriptive reports, any change (0) is the default for other report types.

#### 2.2.9 Report conduct options

In case of larger report series it may be useful to decide whether or not to use already existing results or recalculate them. Additionally, the sensitivity with which the pipeline continues despite of potential flaws may be adapted.

`forcecalc`(int 1) Force new calculations instead of taking existing results. 0=using available results and add only new ones to save computational time, 1=calculate everything new (Default)). ‘0’ should only be chosen when repeating the computations with identical data and unchanged analysis settings.

`breakreport`(int 0) This setting is foremost relevant when generating multiple reports. By setting `breakreport` to 1, any critical incident will terminate computations. By default, `dqrep` continues to compute the remaining reports.

## 3 Technical details

`dqrep` is a wrapper command that triggers a set of about 80 newly created Stata ado and support files to conduct data quality assessments for data sets with unknown issues. It incorporates several existing Stata functions for statistical analysis and graphical display, such as `tabplot, vioplot, coefplot, catplot, xtreg, mixed, xtmelogit,` and `lowess`. As listed above, multiple options control the use of source and target folders, data and metadata files, variable selections, report formatting, and analysis settings. A significant challenge was ensuring robust analysis execution despite deficient data, while maintaining readable output. `dqrep` strives to perform as many data quality checks as possible within the scope defined by the user. Guided by the underlying data quality framework(Schmidt et al., 2021), the `dqrep` pipeline executes four distinct assessment steps:

1. Integrity. This processing stage addresses flaws related to incorrect `dqrep` pipeline specifications, improper formats, unavailable data elements, non-unique records, issues with merging and appending files, as well as data type mismatches. Deficiencies are classified and managed according to three categories: (1) critical issues leading to a program termination or the generation of an error report - immediate forced program termination only occurs at the inital stage in case of significant program call errors; (2) warnings, which indicate that finding are likely to impair correct assessments; (3) notes, which describes shortcomings such as missing metadata information that may limit the scope of a report without impairing the correctness of the assessments. Additionally, `dqrep` recommends actions to remedy detected flaws. Notably, `dqrep` attempts to correct potential errors, such as converting incorrectly provided string values to numeric values, with any modifications reported in a dedicated section of the quality report. Integrity issues are visualized solely through tables.
2. Completeness. In the second step, `dqrep` assesses the number and types of missing values. This includes evaluating missing data from entire observational units (unit missingness) and missing values within observational units (item missingness). Proper assessment of unit missingness relies on the availability of variables that indicate the reasons for missing units. Completeness issues are visualized using tables.
3. Consistency. Consistency checks focus on identifying inadmissible or uncertain values. These checks rely on the existence of appropriate information in the metadata XLSX file, for example admissible ranges for a variable. Consistency issues are visualized using tables and graphs.
4. Accuracy. Finally, `dqrep` targets unexpected distributions and associations, such as observer or device effects, and time trends. Visualizations play a major role. Nonparametric regression models (lowess) are used to visualize time trends, while mixed models estimate the variance attributable to observers, devices, or centres. Various types of graphs, including histograms and violin plots, illustrate distributions. Before analyses are conducted, checks are performed to determine whether certain statistical approaches can be reasonably applied, such as a minimum required number of observations to start computations.

Metrics from the latter three stages can be used to grade the severity of issues such as the variance proportion explained by a device, the number of range violations, or the proportion of missing values for some variable. If requested, these metrics are graded into defined data quality categories and graphically displayed to provide visual guidance on the severity of issues. Grading criteria can be modified using the ancillary *dq_set.ind.xlsx* file.

To place each issue in one problem category, `dqrep` conducts by default selected data cleaning steps after an issue has been encountered. For example, observations outside admissibility ranges are removed before checking for statistical outliers. This approach improves the interpretation of results. However, such data cleaning steps can be suppressed if they are deemed inappropriate within a specific scenario.

### 3.1 Input

`dqrep` uses two types of data as input. First, target data files, which contain the data to be assessed, such as measurements or survey results. `dqrep` can combine multiple target data files into a single analysis file based on a set of specified ID-variables.

The second type of data is a single metadata file that describes the target data and specifies requirements or expectations about these data (Richter et al., 2019). Additionally, the attributes can establish links between variables from the target data to incorporate process information—such as the time of data collection, observer, or devices—into the assessments of measurements. This enables the automated computation of time trends, as well as device or observer effects. The third set of information in the metadata file concerns the role of variables in a report, such as the order in the output, whether a variable should receive a brief (minorvars) or an in-depth analysis (keyvars). More than twenty metadata attributes can be specified but `dqrep` will work with any provided subset.

The use of a metadata file is optional. Certain requirements or expectations can also be provided through the `dqrep` command options. This approach may be convenient for smaller data quality assessment tasks. However, it is only useful for information that remains invariant across the targeted variables. For frequent and repeated assessment tasks, specifying information in a separate .xlsx file is strongly recommended, as this greatly facilitates communication, editing and reuse.

### 3.2 Output

As of Stata 15 or higher, `dqrep` can create PDF or DOCX reports. For illustration purposes, a full PDF report is provided as a supplementary file based on Use Case 4. Figure 1 displays selected output elements. Most report sections can be requested individually by selecting the corresponding `reporttemplate` option. In addition, a structured results log file is available, and HTML dashboards can be requested to inspect graphs.

**Figure 1.**
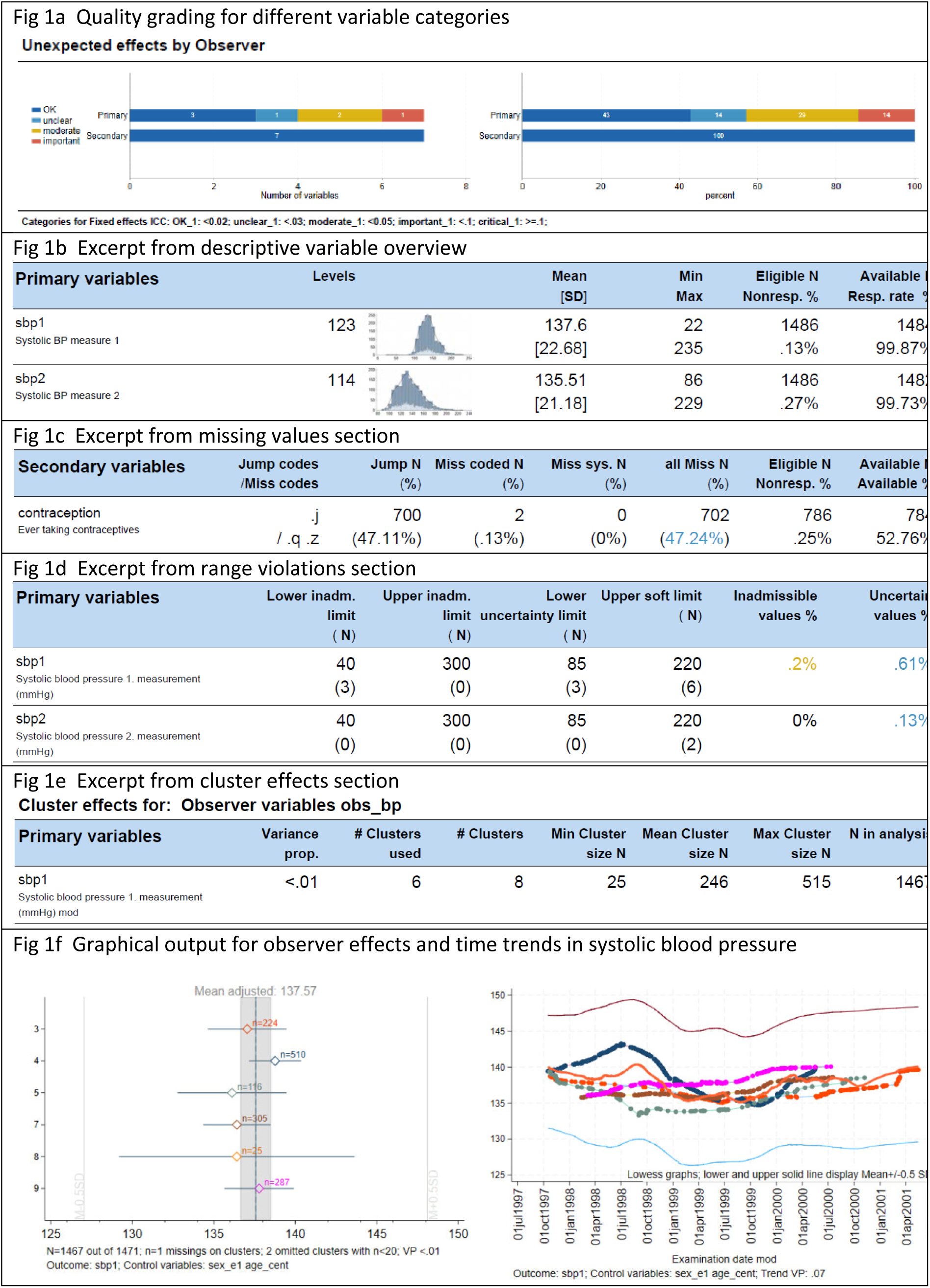
Sample output from dqrep data quality PDF reports.

Per default, only the main report and a log output file are saved. The log output provides more insight on statistical aspects compared to the PDF or DOCX reports. The display language in the PDF or DOCX reports can be changed.

The `store` option enables all additional files generated during the report creation to be retained. This includes graphs, assessment result files for post processing, and modified target data. `store(1)` will include the graphs and Stata .dta files. The applied naming convention for individual level data is ‘reportname’_[data].dta. [data] may either be “targetdata_original” or “targetdata_modified”. “targetdata_original” includes all variables and cases referenced in the report. The only modifications involve the encoding of missing values as specified through the command syntax or metadata file. “targetdata_modified” contains, as implied by the name, additional changes to the values such as recodings of range violations or outliers to missing values and potentially binary recoded categorical variables (suffix *_br). The modified data is only created if at least one such change takes place. In addition to the files listed above, assessment results will be stored based on the naming convention ‘reportname’_results_[resulttype].dta. This file contains all information generated during the assessment process. Whether or not results refer to modified data is indicated by a binary flag in the column “modified”. The rather technical variable names have been chosen to enable the reuse of these files. For example, result files are structurally identical and can be appended across examinations or studies to benchmark findings, using the `benchmark` option. Specifying `store(2)` will additionally store the results as XLSX files. The XLSX file starting with “DQvarhlp” gives a brief explanation of the variables containing the data quality assessment results.

### 3.3 Graphs in dqrep

Most graphs in `dqrep` are non-standard. Examples are provided below:

- Histograms combine both binned and non-binned views, enabling the inspection from both perspectives in a single image. Binning follows an adapted version of Rice’s Rule. It uses the number of value levels instead of the number of observations to decide on the number of bins. By default, no binning takes place when fewer than 30 value levels are present. Information on limits is added to the histograms if provided through metadata.
- Cluster-related graphs visualize the observed effect sizes relative to the overall distribution as this is key for the interpretation in addition to the point estimates and confidence intervals. Therefore, by default, orientation lines or graphs ±0.5 standard deviations are drawn. In the lowess output, this band also visualizes unexpected changes in the standard deviation of the outcome of intrest. In margins plots, clusters without variance in the outcome variable are not excluded from the graph to improve the overview.
- Violin plots encode subgroup size using color intensity, which decreases for subgroups with low counts.

If there is interest only in graphs but not in reports, a convenient way to proceed is requesting the output type of interested, using the `reporttemplate` option, sided by the specification `reportformat(noreport) dashboard(1)`. This will create the images only and an additional html viewer to easily navigate through them.

### 3.4 Terminology

`dqrep` was developed in the context of health studies, where data is routinely collected by different observers and devices, sometimes across multiple study centers. This is reflected in the naming of several input parameters (e.g. `devicevars`), and the default output reads accordingly. However, this may complicate the communication of certain report sections, particularly those related to cluster effects. Therefore, users can override the default cluster-related output terminology by using the options `rename_ob, rename_de, rename_ce`. This possibility is also of importance for readable reports when simply assessing group differences across the levels of categorical variables.

## 4 Setting up dqrep

### 4.1 Installing dqrep

`dqrep` can be installed using the `net install` command:

net from https://packages.qihs.uni-greifswald.de/repository/stata/dqrep

`net install dqrep, replace`

`net get dqrep, replace`

In case of rare issues when downloading, the package files can also be directly downloaded as a zip file from: https://dataquality.qihs.uni-greifswald.de/vignettes.html#STATA to be installed from a local folder.

The following Stata ados must be installed to ensure proper functioning of `dqrep`: `linest, catplot, colorpalette, coefplot, grubbs, robstat,` and `moremata.`

This programming syntax can be used, to do so:

net describe sg100, from(http://www.stata.com/stb/stb47)

`net install sg100`

`ssc install catplot, replace`

net describe gr0075, from(http://www.stata-journal.com/software/sj18-4)

`net install gr0075`

`ssc install coefplot, replace`

`ssc install grubbs, replace`

`ssc install robstat, replace`

`ssc install moremata, replace`

Afterwards, Stata needs to be restarted.

### 4.2 Setting up target data

`dqrep` can be used to assess selected variables from the active dataset. In this case, all information required to assess the data must be provided in the program call. Alternatively, `dqrep` can be applied to one or multiple target data files in a single run. This can be accomplished by specifying the list of relevant target files in the program call using the `targetfiles` option or by listing the file name in the `sourcefilename` column of the XLSX metadata file. When more than one target data file is used, multiple key variables can be specified to ensure a 1:1 match by using the `idvars` option. Variable names must be unique across all included data files, except for variables used for linking the files.

### 4.3 Setting up a metadata XLSX file

Metadata can be provided through the command call. This approach is useful for metadata information that can be used across all targeted variables; otherwise, misleading results will occurr. So, while simple to use, this limitation must always be kept in mind. To overcome this limitation, the full potential of `dqrep` is fully realized when providing a second file, a spreadsheet-type .xlsx metadata file. This file may include labels, missing value codes, value ranges, and links to process variables such as examiners or devices. Only one single metadata file is allowed. The metadata file must adhere to the some formatting rules. Metadata is expected in an Excel .xslx format. This format was chosen to facilitate nontechnical editing of any content. The first row in the file must use column names identical to the lowercase column names listed below; otherwise, any contained information will be ignored. Only the `var_name` column is obligatory, other columns can be provided as needed. Admissible columns are:

`var_name` The variable name must match exactly with the name of the corresponding variable in the target data files. Variable names must be unique.

`varlabel` The label of the variable. This can be a longer description that conveys the meaning of the variable as clearly as possible. This label is primarily used for table outputs.

`varshortlabel` The shortened version of the variable label. The primary purpose of this alternative output is to improve readability, especially in graphs.

`value_label` Contains the value labels of categorical variables. This can be formatted either in STATA syntax (e.g., 0 “no” 1 “yes” 2 “don’t know") or using a pipe symbol to separate levels “0=no | 1=yes | 2=don’t know”.

`data_type` Specifies the data type of the variable. Four data types are recognized: string: for character or string variables; integer: for variables containing whole numbers; float: for variables with decimal values; datetime: for variables in a date / time formats. Any other entries will be ignored. The data type is used for data integrity checks and to determine analyses of relevance.

`scalelevel` Specifies the scale level of the variable, based on the Stevens classification. The recognized values are ’nominal’, ’ordinal’, ’interval’, and ’ratio’. For detailed explanations, refer to the relevant literature. This attribute controls the selection of applicable statistics, such as outlier checks.

`missinglist` A list of values (numeric or Stata missing codes) that represent missing data. (Example data field entry 998 999 .x.z). When providing numerical missing codes, missings are encoded before analyses.

`jumplist` A list of values (numeric or Stata missing codes) that represent omitted data. Jumps refer to data points that were not collected due to the data collection design. (Example data field entry: 888 .j). When providing numerical jump codes, these are encoded before analyses.

`refcat` Used to recode categorical variables to binary. refcat contains a numlist, optionally including also Stata missing codes, to define the reference category (coded as 0). (Example data field entry in column ’refcat’: 0 1 2)

`eventcat` Used to recode categorical variables to binary. eventcat contains a numlist, optionally including also Stata missing codes, to define the event category (coded as 1). (Example data field entry in column ’refcat’: 3 4 5 6 7)

`limit_hard_low` The lower bound of an inadmissibility limit. (Example data field entry in column ’limit_hard_low’: >0). Breaking this limit is a data error.

`limit_hard_up` The upper bound of an inadmissibility limit. (Example data field entry in column ’limit_hard_up’: <300). Breaking this limit is a data error.

`limit_soft_low` The lower bound of an uncertainty limit. (Example data field entry in column ’limit_soft_low’: >=50). Breaking this limit may be an indication for a data error.

`limit_soft_up` The upper bound of an uncertainty limit. (Example data field entry in column ’limit_soft_up’: <=150). Breaking this limit may be an indication for a data error.

`key_observer` Links the name of the variable representing observers/examiners, who collected the data in `var_name`. This information is used to assess observer effects. Applicable variable names must be entered in this column. Multiple variables can be specified, separated by spaces. `key_device` Links the name of the variable representing the devices used to collect values in `var_name.` Based on this link device effects can be assessed. Multiple variables can be specified, separated by spaces.

`key_center` Links the name of the centers where the values of `var_name` have been collected.

Based on this link device effects can be assessed. One center variable may be specified.

`key_datetime` Links a date time variable to `var_name` to assess time trends.

`variablerole` Assigns a role to the variable `var_name` for the report. The specified strings corresponds to the following options: `keyvars`, `minorvars`, `processvars`,

controlvars, observervars, devicevars, centervars, timevars, idvars. Please refer to the explanations above.

`var_order` Specifies the order of variables in the report. Integer input is expected, and variables are sorted accordingly.

`sourcefilename` Specifies the name of the Stata .dta file containing *var_name*. This is useful for reports requiring the merging of multiple data files. If a metadata file is provided but no target data file is specified using the *targetfiles* option, *dqrep* assumes the presence of a *sourcefilename* column. If no such column is found, program execution stops.

In addition, the metadata file may contain attributes with custom names, specified via a command option:

`segmentname` Refers to the variable that identifies the segment to which a variable belongs. A segment may represent for example an examination. This column must be specified to generate multiple reports with a single `dqrep` call.

`varselect` Specifies a variable in a metadatafile that defines, which variables is to be included in a report. Specifying ’1’ means that the variable is to be included. Any other code is ignored.

Metadata file specifications and command syntax options can be combined. In cases where both are used, command option settings on program execution override information in the metadata file.

An example of a metadata file is the ancillary file example_metadata.xlsx.

### 4.4 Data quality grading

`dqrep` can grade data quality for selected results. The current scope reflects requirements during data monitoring in observational studies. Gradable indicators relate to completeness, range violations, univariate outliers, variance proportions attributed to cluster level variables (e.g. examiners, devices, centers), and time trends. Default grading rules are provided with the package; however, any grading is highly context specific, and users may wish to adapt rules or skip over any grading entirely.

There are three main approaches to control a data quality grading. Users exert control over whether or not to conduct a grading at all, which indicators to grade, and how to perform the grading. These approaches are listed below:

1. Using the `view_dqi` option, data quality grading can be switched on or off entirely. The two other approaches are only relevant if `view_dqi` is set to “1”. Using a “standard” or “extended” `reporttemplate` enables data quality grading by default with no need to use `view_dqi`.
2. The `select_dqi` option allows individual data quality indicators to be selected for grading. Details are described in the respective option. `select_dqi` will only work in conjunction with a user specified grading file.
3. An XLSX file containing grading rules can be edited as needed and loaded using the `gradingfile` option. It is recommended to base modifications on the provided “example_grading.xlsx” ancillary file. The settings in this XLSX resemble the default `dqrep` grading settings. The first sheet of the file includes instructions on how to modify content. In brief, the following aspect can be adjusted for each indicator: First, the use of up to seven data quality level for each indicator. These levels are: 1 “OK”; 2 “uncertain”; 3 “Note” (used as a contrast to 1 “OK” to provide a note on potential issues, specifying 3 will exclude 4 and above); 4 “low”; 5 “moderate”; 6 “important”; 7 “critical”. Second, logical rules to determine the display of a given category. Third, the color associated with each data quality level to display in the report. Currently, the same rule set is applied to all variables in a report.

## 5 Examples

The following examples use anonymized data from SHIP (Volzke et al., 2022) to illustrate selected uses of `dqrep`. These files are also included as ancillary files with the package. The data set (N=1,486) comprises selected somatometric and blood pressure variables, laboratory markers, and variables from a medical interview. Issues have been added to provoke certain quality findings for educational purposes but also to illustrate potential pitfalls of the pipeline when ignoring more advanced setting options. For simplicity, all examples below assume that the working directory is set to the folder containing the target data and metadata files (example_targetdata.dta, example_metadata.xlsx) as follows:

`cd “your working directory containing example_targetdata.dta/ example_metadata.xlsx"`

### 5.1 Example 1

Creating a descriptive overview of all variables in the active data set.

`use example_targetdata.dta, clear`

`dqrep`

Simply typing dqrep suffices. By default, if nothing is specified, all results are written into a subfolder named “*DQ-resultfolder”.* The output highlights a potentially insufficient handling of missing data that can be remedied as follows:

### 5.2 Example 2

Creating an extended descriptive overview of all variables in the active data set with an improved handling of missing data as a DOCX file.

`use example_targetdata.dta, clear`

`dqrep, targetfiles("example_targetdata") rd(Example2) ///`

`itemmisslist(999,.d,.t,.v,.z) itemjumplist(.j) ///`

`reportname(Example_2) reporttemplate(D3) reportformat(docx)`

A report name has now been assigned to avoid overwriting the previous report. In addition, a DOCX report has been requested with an extended descriptive overview using the D3 reporttemplate. Results are stored as well in the subfolder *DQ-Example2*.

### 5.3 Example 3

Creating a full PDF data quality report with extensive metadata specified directly via command syntax, retaining the key created result data files.

`dqrep, targetfiles("example_targetdata") rd(Example3) ///`

`itemmisslist(999,.d,.q,.t,.v,.z) itemjumplist(.j) ///`

`reportname("Example_3") reporttitle("Example 3 Standard data quality report") ///`

`reporttemplate(standard) reportformat(pdf) dashboard(1) ///`

`keyvars("sbp1 sbp2 dbp1 dbp2") ///`

`minorvars(cholesterol ldl stroke diab_known waist contraception) ///`

`observervars(obs_bp) devicevars(dev_bp) ///`

`controlvars(age sex) idvars(id) timevars("exdate")`

This example shows how to conduct already an extensive data quality report. The options used include missing value codes (`itemmisslist`), permitted jump codes (`itemjumplist`), report names, and report title. This report focuses on blood-pressure related outcomes (`keyvars`), requesting a single report page for each of the listed variables. For other variables, specified as `minorvars`, only table outputs are created. Additionally, the examiners (`observervars`), and devices (`devicevars`) are identified, as well as variables to control for when checking these effects (`controlvars`). An id variable is also defined (`idvars`).

### 5.4 Example 4

Creating a full PDF data quality report using a XLSX metadata spreadsheet with an additional dashboard.

`dqrep, rd(Example4) reportname(Example_4) metadatafile("example_metadata.xlsx") ///`

`reporttitle("Example 4 Extended data quality report based on a metadatafile") ///`

`reporttemplate(extended+) dashboard(1) gradingfile(example_grading.xlsx)`

While shorter, this command is much more powerful because the spreadsheet enables a much finer-grained provision of metadata to control the report generation. The key difference from the previous example is that all attributes can vary at the level of individual variables. For instance, distinct process variables can be assigned to each variable in the report and limits for range checks can be applied. An HTML dashboard is additionally requested to browse images. This will automatically retain result files in the subfolder Example4.

### 5.5 Example 5

Creating multiple PDF data quality reports with an added benchmarking of data quality issues

`dqrep, rd(Example5) reportname(Example_5) metadatafile("example_metadata.xlsx") ///`

`reporttitle("Example_5 Data quality multi report") ///`

`reporttemplate(standard) segmentname(segments) benchmark(2) store(2)`

In addition to the previous command, the `segmentname` option is used to specify the name of a variable that identifies the distinct underlying examinations. For each segment, a distinct report is created. While only of illustrational purpose here, this possibility is essential for handling large data collections. To compare the data quality across reports, the `benchmark` option is applied. The value provided through this option is used to include only variables with a detected minimum problem level in the report according to the rules specified in the grading level Excel sheet `dq_set_ind.xlsx`.

The specifications in the current report result in four separate data quality reports, accompanied by a benchmarking report overview that contains only variables with at least a moderate level of issues.

Many more options exist to adapt data checks and data quality reporting to individual needs.

## 6 Conclusions

The `dqrep` package provides a broad range of functionalities for assessing data properties and quality, from simple descriptive overviews to comprehensive data quality reports. There are some limitations, however. The focus of `dqrep` is on an accessible formalized workflow to maximize the repeatability and comparability of results across assessments rather than interactive assessments. `dqrep` takes a standardized approach to assess data properties and data quality. As such, it neither intends to nor replaces any highly sophisticated tailored statistical analyses. It remains the user’s responsibility to infer the underlying reasons for any observed findings and to make decisions on additional in depth analyses. Furthermore, figures require manual inspection to detect issues not accounted for by the provided metrics. Due to a large chain of analytical decisions, the pipeline may produce misleading results, particularly in the absence of carefully curated metadata. In fact, wrong metadata has proven to be a key issue in complex application scenarios of this tool. `dqrep` has been designed to run robustly with deficient data, trying to avoid uncontrolled program terminations. Yet, using it with new datasets may unveil yet undiscovered pipeline issues.

`dqrep` currently focuses solely on numerical variables and does not support string variables. Including string variables would considerably expand the application scope. Another extension will be the analysis of repeated measurements. `dqrep` enables a grading of selected data-quality metrics into distinct issue severity categories. Any such grading is highly context specific and users are advised to revise and adapt the rules to their specific needs as outlined above.

`dqrep` is not made for big data. Less than 500 variables should be included in single reports, but results from single reports may be combined into larger reports. Be aware that large case or variable numbers may result in very long computational times.

Extensive use of information from Excel metadata files to control data quality assessments, combined with numerous options for customizing reports, enables large-scale applications. `dqrep` is used in major cohort studies such as SHIP (Volzke et al., 2022), and the GNC (German National Cohort, 2014), For example, a single `dqrep` call is used in the current routine data monitoring reporting pipeline within SHIP to assess more than 4000 variables across 60 reporting segments with a subsequent benchmarking. Creating extensive metadata information for such purposes is time-consuming. Yet, having metadata available in spreadsheet format not only broadens the scope of automatable assessments. It also enhances transparency by providing accessible machine-readable rules against which the data is checked. This increases the FAIRness (Wilkinson et al., 2016) of data quality assessments and their results. Data quality reports, as provided by `dqrep,` could serve as valuable supplements to research articles, providing deeper insights into the data underlying substantive scientific analyses.

## 7 Supplementary data

Sample data and sample code are available as ancillary files in the **dqrep** Stata package. A report based on Example 4 is available as a supplementary file.

## 8 Availability of data and materials

All datasets analyzed are provided with the `dqrep` package as ancillary files.

## Data Availability

This work does not contain original data but is a guidance work on appropriately assessing data

https://dataquality.qihs.uni-greifswald.de/vignettes.html#STATA

## Acknowledgements

This work was partially supported by the German Research Foundation (DFG: SCHM 2744/3-4, NNFDI4Health project (www.nfdi4health.de) – Project Number 442326535, by the European Union’s Horizon 2020 research and innovation programme under grant agreement No 825903 (euCanSHare project), and the German National Cohort (NAKO) as funded by the Federal Ministry of Education and Research (BMBF: 01ER1301A and 01ER1801A). ChatGPT has been used for language editing and the development of the HTML dashboard.

I extend my gratitude to the SHIP team members, especially Dr. Birgit Schauer and Dr. Janka Schössow, for their efforts in setting up metadata and their support in running `dqrep` on multiple datasets to provide feedback on its functionality since 2019. Additionally, Stephan Struckmann offered valuable assistance with the data quality webpage.

Two existing Stata packages (tabplot and vioplot) have been integrated into dqrep with minor adjustments to enhance their operation within the dqrep data quality assessment workflow. Full credit remains with their respective authors.

## About the author

Carsten Oliver Schmidt is full professor at the University Medicine Greifswald, leading the functional division “Quality in the Health Sciences”. His work focuses on population-based epidemiology, incidental MRI findings, statistics, research methods, research infrastructures, standards, and data quality.

